# IFNβ-1b treatment leads to changes in the B cell subset and cytokine secretion profile in patients with relapsing-remitting multiple sclerosis

**DOI:** 10.1101/2022.02.25.22270266

**Authors:** Daniel W. Mielcarz, Alan J. Bergeron, John K. DeLong, Alexandra Dias, Kathleen M. Smith, Karen L. Mack, Lloyd H. Kasper, Jacqueline Y. Channon

## Abstract

In order to explore the effect of IFNβ-1b treatment on B cell phenotype and function in RRMS patients, blood was drawn from RRMS patients before treatment and again 2 and 6 months after every other day injections of IFNβ-1b. Cryopreserved peripheral blood mononuclear cells (PBMCs) were thawed and stained with panels of antibodies against B cell surface antigens and the intracellular cytokines, IL-10 and IL-6. At baseline, PBMCs from RRMS patients have increased frequencies of B1 cells and a decreased frequencies of memory B cells when compared with PBMCs from age- and gender-matched healthy controls. CpG-stimulated PBMCs from patients treated with IFNβ-1b show an increase in IL-10 production and a decrease in IL-6 production by naïve, memory and B1 cells compared with healthy controls. In addition, this treatment alters the composition of circulating B cell subsets, leading to an increase after six months in circulating naïve B cells and a decrease in both memory and B1 cells, both cell types of which are potentially pathogenic in RRMS. Patients were divided into two groups based on disease activity, no/low or moderate/high. A significantly higher frequency of B1 cells and higher expression of CD27 on these cells was seen at baseline in the moderate/high disease activity group to IFNβ-1b compared with patients in the no/low disease activity group. Although the number of subjects in this study was limited, these findings suggest that alterations in the B cell compartment may be a mechanism by which IFNβ-1b reduces disease activity.

## Introduction

Multiple sclerosis is a chronic inflammatory disease of the CNS leading to demyelination and neurodegeneration (1). Relapsing-remitting multiple sclerosis (RRMS) is associated with acute inflammatory episodes resulting in a reduction in neurological function. T_h_1 and T_h_17 subsets of myelin-specific CD4+ cells are thought to be the main pathogenic cells in RRMS (2). However, recent studies provide strong evidence for a role for B cells in the pathology of RRMS (3). Therapy directed at the depletion of CD20+ B cells has been shown in Phase II clinical trials to be effective in reducing relapse rate and reducing lesions as detected by MRI in those with RRMS (4, 5). However, a Phase II trial with atacicept, a soluble inhibitor of B cell survival, showed higher levels of inflammation and MRI activity in treated patients, indicating that there may be a pleiotropic effect of B cells in RRMS (6). It is known that CSF samples from patients with active RRMS are enriched in memory B cells and that persisting plasmablasts are a major source of antibody-secreting cells responsible for oligoclonal bands in the CSF (7). In addition to oligoclonal bands found in the CSF, MS patients have also been shown to have a unique autoantibody signature in their serum (8).

Mature human B cells are typically divided into two subsets, naïve (CD27-) and memory (CD27+) (9). A subset of human autoantibody producing cells, B1 cells (well known in the mouse model), has recently been described (10). B1 cells present antigen efficiently, produce autoantibody and can induce naive CD4 T cells to become T_h_17 cells, a major pro-inflammatory compartment that is involved in the pathogenesis of human RRMS (11). IgM autoantibody produced by B1 cells can function to clean up self-antigen from dead and apoptotic cells (12). Human B1 cells are found in low frequency in peripheral blood and are CD20+CD27+CD43+ (10).

IL-10 is a potent anti-inflammatory cytokine produced primarily by T_h_2 and T_reg_ cells, although it can be made by most leukocytes (13). IL-10 production by B cells has been associated with regulatory, anti-inflammatory functions. Specifically, it has been demonstrated to be important for recovery from EAE, the animal model of MS (14-17). IL-10 secretion by B cells stimulated with CD40 ligand has been found to be decreased in MS patients when compared with healthy controls, indicating a potential defect in production of this cytokine by B cells in MS patients (9). IL-6 is a pro-inflammatory cytokine produced primarily by T cells and macrophages (18). IL-6 is important for the differentiation of T_h_17 cells, as well as the differentiation and proliferation of B cells (19, 20). IL-6 production by B cells may potentially be a signal of a pro-inflammatory phenotype (21).

While recent studies have begun to explore the role for B cells in RRMS, little is known about the effect of IFNβ-1b on the composition and function of B cell subsets and their regulatory markers in peripheral blood during disease (3). This study aims to clarify that effect by examining the frequency of B cells subsets and the cytokines produced by B cells isolated from RRMS patients pre-treatment, and two and six months after initiation of treatment.

## Methods

### Patients

For this trial, 10 patients with clinically defined RRMS (McDonald criteria) were enrolled and treated with IFNβ-1b every other day (qOD). The trial was approved by the Geisel School of Medicine IRB. The treatment window was 6 months during which time 200 ml blood was drawn and assayed for immunologic changes at baseline, month 2 and month 6 post-therapy. The principal immunologic outcome was the effect of therapy on B cells as determined by phenotypic changes and intracellular cytokine expression using cryopreserved PBMCs. Patients were divided into two groups based on these criteria: (i) no relapse within 1 year while on IFNβ-1b, (ii) EDSS score: stable or a maximum increase of 1.0 within 1 year while on IFNβ-1b, (iii) no new enhancing lesions by MRI within 1 year while on IFNβ-1b. There were 6 patients with no/low disease activity and 4 with moderate/high disease activity (see Table 1).

**Table 1.** Patient demographics and clinical response to IFNβ-1b.

| **Patient**<br>**(gender/age**<br>**range)** | **Clinical response**<br>**to IFNβ-1b** |
| --- | --- |
| F, 30-39 | Yes |
| M, 40-49 | Yes |
| F, 40-49 | Yes |
| F, 40-49 | Yes |
| M, 50-59 | Yes |
| F, 50-59 | Yes |
| F, 40-49 | No; relapse |
| M, 20-29 | No; new lesions on<br>MRI |
| F, 50-59 | No; new lesions on<br>MRI |
| M, 50-59 | No; became<br>secondary<br>progressive |

### PBMC Isolation and cryopreservation

Blood was collected in 200ml blood bags containing sodium citrate. Peripheral blood mononuclear cells were isolated via density gradient separation. Briefly, 20 ml of blood was overlaid with 20 ml of Histopaque-1077. Blood was centrifuged at 1800 rpm for 30 minutes at 25°C. Following centrifugation, PBMCs were removed and washed 2 times via centrifugation with HBSS without calcium and magnesium ions. PBMCs were counted and adjusted to desired concentration in human AB serum + 10% DMSO. PBMCs were then frozen overnight at -80°C in a cryopreservation chamber, then transferred to -140°C for long-term storage.

### Flow cytometry for B cell subset analysis

A violet amine viability dye (Invitrogen) and anti-CD14-Pacific blue was present in all panels as a dump channel and viability after thawing and washing was >80% of thawed cells. Antibodies were from BD (CD43 FITC) or from BioLegend and were all titrated before use (see Table 2). Each panel contained duplicate fully-stained samples and a fluorescence-minus-one (FMO) tube for each antibody. Patient PBMCs for all time points (27 samples) were thawed on the same day and stained in 96-well plates. Healthy controls (10 samples) were thawed on a different day and stained in the same way as for patient samples. Inter-day staining variability was controlled for by use of an aliquot of cryopreserved PBMCs from the same donor that was thawed and stained daily in parallel with patient or healthy control PBMCs. Flow cytometry was carried out using a 7-color MACSQuant cytometer (Miltenyi Biotec). FlowJo software (Tree Star, Inc.) was used for post-hoc compensation and analysis.

**Table 2.** Antibodies used for surface staining panel.

| **Epitope** | **Panel** | **Clone** | **Supplier** |
| --- | --- | --- | --- |
| CD43 | FITC | 1G10 | BD Biosciences |
| CD70 | PE | Ki-24 | BD Biosciences |
| IgD | PerCP-Cy5.5 | IA6-2 | BD Biosciences |
| CD5 | PE-Cy7 | UCHT2 | BioLegend |
| CD27 | APC | MT271 | BioLegend |
| CD20 | APC-eF780 | 2H7 | eBioscience |
| CD14 | VioGreen | TUK4 | Miltenyi |

### In vitro stimulation and intracellular cytokine staining

Intracellular staining for IL-10 and IL-6 was performed as a modified version of a previously described procedure (22). Cryopreserved and thawed PBMC (for both patients and healthy controls) were cultured at 5×10^6^/ml in 12-well, flat-bottom plates in AIM V medium supplemented with 10% pooled human AB serum for 18 hours with 3 µg/ml CpG oligodeoxynucleotide type B-2006-G5 (InvivoGen, CA) at 37^°^C in a humidified atmosphere containing 5% CO_2_. PMA (50 ng/ml final concentration; Calbiochem) and ionomycin (750 ng/ml final concentration; Calbiochem) were added after 12 hours and Brefeldin A and monensin (3 µg/ml and 2 LM final concentration, respectively; BioLegend) were added after 14 hours. PBMCs were stained in V-bottom plates for viability (yellow amine viability dye; Invitrogen) for 30 min prior to surface epitope and intracellular cytokine staining. Cells were stained for the surface epitopes in Table III with 100 µl cocktail containing all antibodies (see Table 3) plus 1.25 mg/ml Cohn fraction IgG block (Sigma) in PBS and the protein transport inhibitors brefeldin A and monensisn (PTI). Cells were incubated with surface marker stains for 30 min with shaking at 4^°^C. Cells were washed twice in PBS with protein transport inhibitors and fixed overnight with 0.5% EM grade p-formaldehyde (Polyscience). On the following day, fixed cells were pelleted and 200 Ll fix/perm reagent (BD Biosciences) was added to each well to prepare the cells for intracellular staining. After 30 min in fix/perm buffer at room temperature cells were washed twice with permeabilization reagent, then stained with anti-cytokine antibodies or isotype control antibodies in permeabilization buffer with shaking for 30 min at room temperature. Cells were then washed twice with permeabilization reagent and resuspended in 200 µl PBS for flow cytometry using a 3-laser, 10-color Gallios flow cytometer (Beckman Coulter).

**Table 3.** Antibodies used for Intracellular Cytokine Staining Panel.

| **Epitope** | **Fluorochrome** | **Clone** | **Supplier** |
| --- | --- | --- | --- |
| CD43 | FITC | 1G10 | BD |
| CD27 | PE-CF594 | MT271 | BD |
| CD24 | PerCP-eFluor® 710 | SN3 A5-2H10 | eBioscience |
| CD19 | PE-Cy7 | H1B19 | Biolegend |
| CD20 | Alexa Fluor 700 | 2H7 | Biolegend |
| CD69 | APC-Cy7 | FN50 | Biolegend |
| CD5 | Brilliant Violet 421 | UCHT2 | Biolegend |
| CD14 | VioGreen | TUK4 | Miltenyi |
| IL-6 | APC | MQ2-13A5 | Biolegend |
| IL-10 | PE | JES3-9D7 | Miltenyi |
| Rat IgG1 | APC | RTK2071 | Biolegend |
| Rat IgG1 | PE | RTK2071 | Biolegend |

### Statistical analysis

Groups were compared using Prism software (GraphPad). RRMS patient timepoints and healthy controls were compared using One-way ANOVA with Tukey’s multiple comparison post test. Changes over time following treatment were compared using repeated measures ANOVA with Tukey’s multiple comparison post test.

## Results

### B cell subset changes following IFN-β1b treatment

Circulating B cells that have not yet differentiated into plasmablasts have traditionally been divided into two subsets: naïve B (CD19+CD20+CD27-) and memory B (CD19+CD20+CD27+). Compared to naïve B cells, memory B cells are characterized by a large increased expression of CD27, with small increases in CD19, CD20, CD69, CD5 and SSC expression, whereas FSC and expression remain unchanged. While long described in mice, only recently has a phenotype for a third subset, B1 B cells, been described in humans(10, 23). These cells are CD19+CD20+CD27+CD43+, while memory B cells are CD19+CD20+CD27+CD43-. B1 B cells are somewhat larger, activated cells than naïve and memory B cells characterized by a large increase in expression of CD43 and CD5, with a small increase in expression of CD27 and slightly decreased expression of CD19 and CD20.

To determine the level of these subsets in RRMS patients treated with IFN-β1b, cryopreserved PBMC from patients and healthy controls were thawed and stained for CD20, CD27 and CD43. To allay concerns that the cryopreservation process may change B cell subset frequency, 5 healthy controls were examined before and after freezing. Naïve and memory B cells showed stable frequency with a slight but consistent decrease in B1 B cell frequency (Supplemental Figure 1). Cells were gated as illustrated in Figure 1A. Briefly, viable CD14-negative cells were gated, and then CD20 positive B cells. The B cells were further subdivided into three subsets: naïve B (CD27-CD43-), B memory (CD27+CD43-) and B1 (CD27+CD43+). Subset frequencies across all time points and for healthy controls are compiled in Figure 1B. Interestingly, compared to healthy controls, RRMS patients had a significantly higher frequency of B1 B cells at baseline that approached healthy control levels with treatment. Furthermore, at all time points MS patients showed a significantly lower frequency of memory B cells when compared with healthy controls. No significant difference in naïve B cell frequency was seen, however, a trend towards an increase in naïve cells that was concomitant with the decreases in the other two B cell subsets is evident. In Figure 1C B cell subset frequencies in individual patients are plotted. Here, significant (p<0.05) decreases in both B1 and memory B cell frequencies are seen, along with a significant (p<0.001) increase in naïve B cell frequency. Patients with no/low disease activity up to one year following the beginning of treatment to IFN-β1b are plotted in red, and moderate/high disease activity in black, but clinical response does not seem to affect either the initial level of subset frequency or response to treatment. Taken together, these results suggest a significant modification of the circulating B cell population in RRMS that can be altered with IFN-β1b treatment.

**Figure 1.**
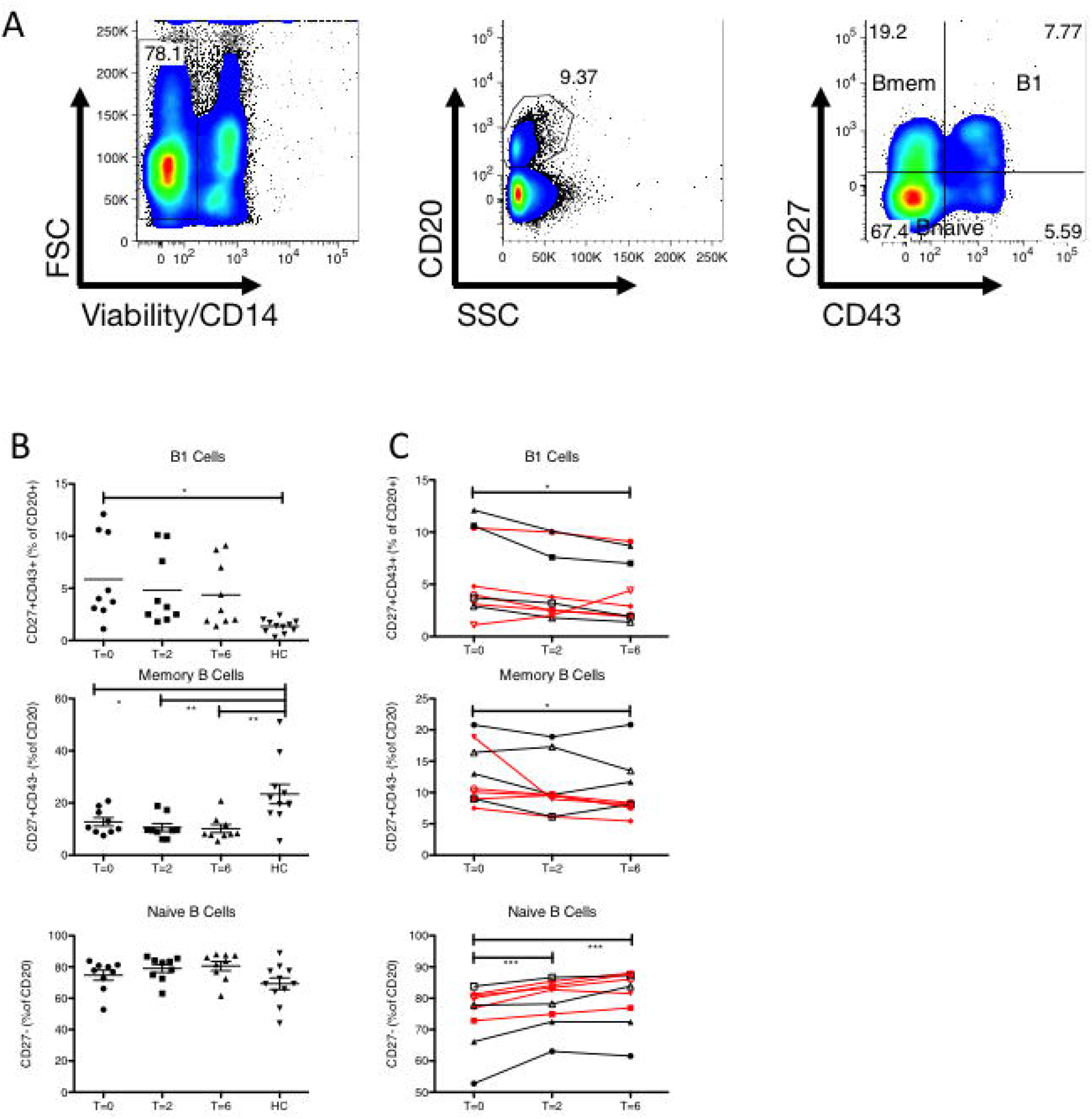
B cell subset frequencies differ in RRMS patients and healthy controls, and change over time with IFN-β1b treatment. (A) Gating strategy for delineating subsets. From left to right: viable, CD14-cells were gated, then CD20+ B cells. In the far left bivariate plot, CD27 and CD43 expression was used to delineate naïve B cells (CD27-CD43-), memory B cells (CD27+CD43-) and B1 B cells (CD27+CD43+). Representative data from 1 RRMS patient at baseline is shown. (B) Compiled subset frequencies from 10 RRMS patients at baseline, 2 and 6 months following initiation of IFN-β1b treatment and 10 healthy controls. Each data point represents the average of three separate samples. The horizontal bar indicates the mean frequency for each time point, and error bars represent the standard error of the mean. Data analyzed by one-way ANOVA with Tukey’s multiple comparison post-test. * p<0.05, **p<0.01. (C) Subset frequencies for individual RRMS patients following initiation of IFN-β1b treatment. Clinical responders to IFN-β1b are shown in red. Each data point represents the average of three separate samples. Data analyzed by repeated measures ANOVA with Dunnett’s multiple comparison post-test. * p<0.05, **p<0.01 ***p<0.001.

### Intracellular IL-6 and IL-10 staining of PBMC

PBMC isolated from RRMS patients and healthy controls were stimulated *in vitro* with CpG and stained as described in the materials and methods. Cells were acquired on a flow cytometer and the gating strategy is illustrated in Figure 2. First, a broad PBMC gate was drawn based on forward and side scatter. Second, a gate was drawn to eliminate both dead cells and CD14+ monocytes. B cells were then gated based on CD19 and CD20 expression, and these B cells were further subdivided into three subsets: naïve B (CD27-CD43-), B memory (CD27+CD43-) and B1 (CD27+CD43+). These subsets were then analyzed separately for IL-6 and IL-10 expression. Quadrants were set based on isotype controls.

**Figure 2.**
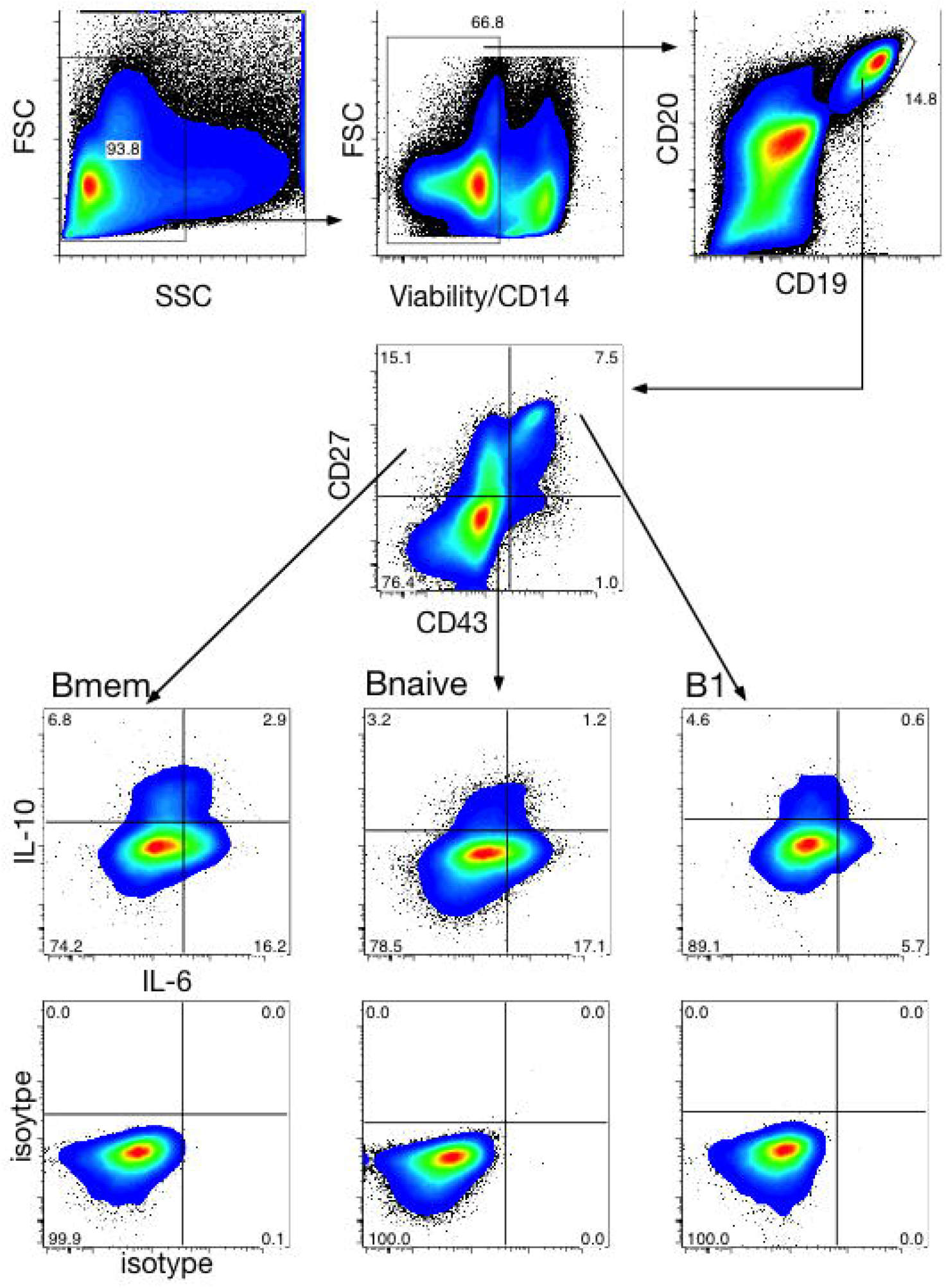
Intracellular cytokine staining of B cells from RRMS patients. Cryopreserved and thawed PBMC were ml in flat-bottom plates in AIM V medium with 10% human AB serum for 18 hours with cultured at 5×10^6^/ 3 µg/ml CpG-ODN type B-2006-G5 at 37^°^C, 5% CO_2_. PMA (50 ng/ml) and ionomycin (750 ng/ml) were added after 12 hours. Brefeldin and monensin (PTI)(3 µg/ml and 2 µM, respectively) were added after 14 hours. PBMCs were stained for viability (yellow amine dye) prior to surface epitope and intracellular cytokine staining. Cytokine quadrants were set based on isotype controls.

Stimulated cells were also examined for CD69 expression, an early activation marker that is constitutively elevated on B1 B cells. As expected (data not shown) after *in vitro* stimulation, nearly all B1 B cells from healthy controls and patients at all three time points were CD69+. Memory B cells had a lower frequency of expression of CD69 following stimulation, and naïve B cells had the lowest expression. CD69 levels following stimulation are not significantly affected by IFN-β1b treatment or by disease status.

### Frequency of IL-6 and IL-10 expression by naïve B cells

Individual samples were analyzed in triplicate for IL-6 and IL-10 expression after culture with CpG and PMA/Ionomycin. Cytokine gates seen in Figure 2 were set based on isotype controls in which the anti-cytokine antibody was omitted from the staining panel – positive cells are those in which the fluorescence intensity exceeds the background staining in the isotype control. The frequency of IL-10 and IL-6 expressing naïve B cells (CD27-CD43-) for all patient time points and healthy controls can be seen in Figure 3A. MS patients treated with IFN-β1b had a significantly higher frequency of IL-10 producing and IL-10 and IL-6 double producing B cells than healthy controls at both 2 months and 6 months following initiation of treatment (p<0.05). Interestingly, MS patients had a significantly higher level of the inflammatory cytokine IL-6 producing naïve B cells at baseline versus healthy controls (p<0.01), and this significant difference continued throughout treatment, although a trend toward a decrease was seen at 6 months following initiation of treatment

**Figure 3.**
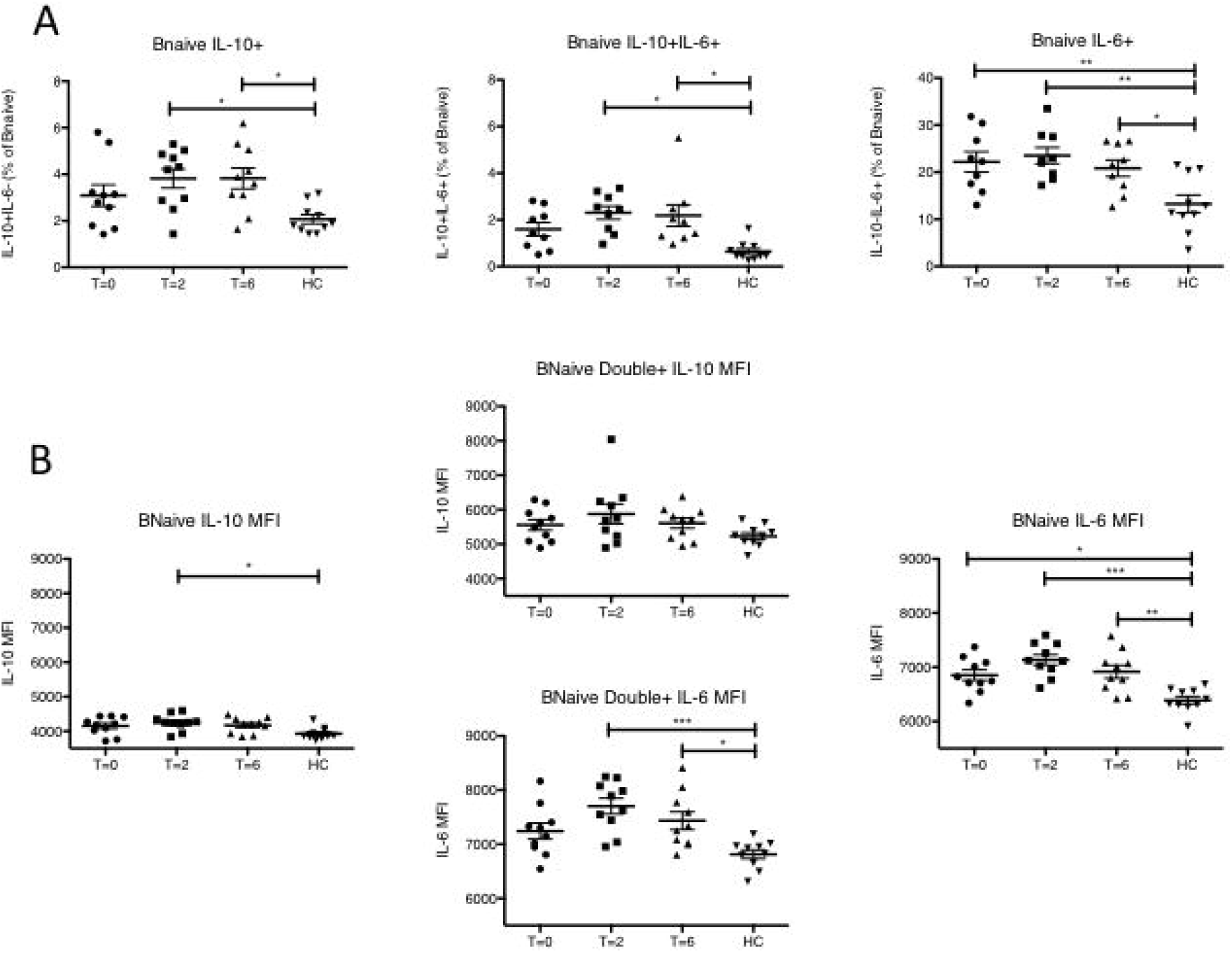
IL-6 and IL-10 expression by naïve B cells differs in RRMS patients and healthy controls and is modulated with IFN-β1b treatment. Cells were analyzed via flow cytometry as shown in figure 2. Each data point is representative of the average of two replicates. The horizontal bar indicates the mean frequency for each time point, and error bars represent the standard error of the mean. Data analyzed by one-way ANOVA with Tukey’s multiple comparison post-test. * p<0.05, **p<0.01, ***p<0.001. (A) Frequency of cytokine positive cells naïve B cells. (B) Geometric mean fluorescence intensity of cytokine positive cells.

Figure 3B presents the level of cytokine expression as measured by geometric mean fluorescence intensity in naïve B cells for healthy controls and patients at baseline and two and six months following treatment. IL-10 levels in single positive IL-10 producing cells were highest 2 months following initiation of treatment, and significantly higher than those of healthy controls. In IL-10/IL-6 double positive cells, IL-6 levels are significantly higher at 2 and 6 months following treatment initiation compared with healthy controls, and IL-10 levels are higher than those of single IL-10-positive cells. Expression levels of IL-6 in IL-6 single-positive cells are significantly higher than healthy controls at all time points, including baseline.

### Frequency of IL-10 and IL-6 expression by memory B cells

The frequency of IL-10 and IL-6 expressing memory B (CD27+CD43-) cells for all patient time points and healthy controls can be seen in Figure 4A. MS patients treated with IFN-β1b had a significantly higher frequency of IL-10 producing and IL-10 and IL-6 double producing B cells than healthy controls at both 2 months and 6 months following initiation of treatment (p<0.05). Unlike in the naïve B cell population, there were no significant differences in frequency of single-positive IL-6 producing cells between MS patients and healthy controls in the memory B cell population, although the mean frequency was elevated at all time points.

**Figure 4.**
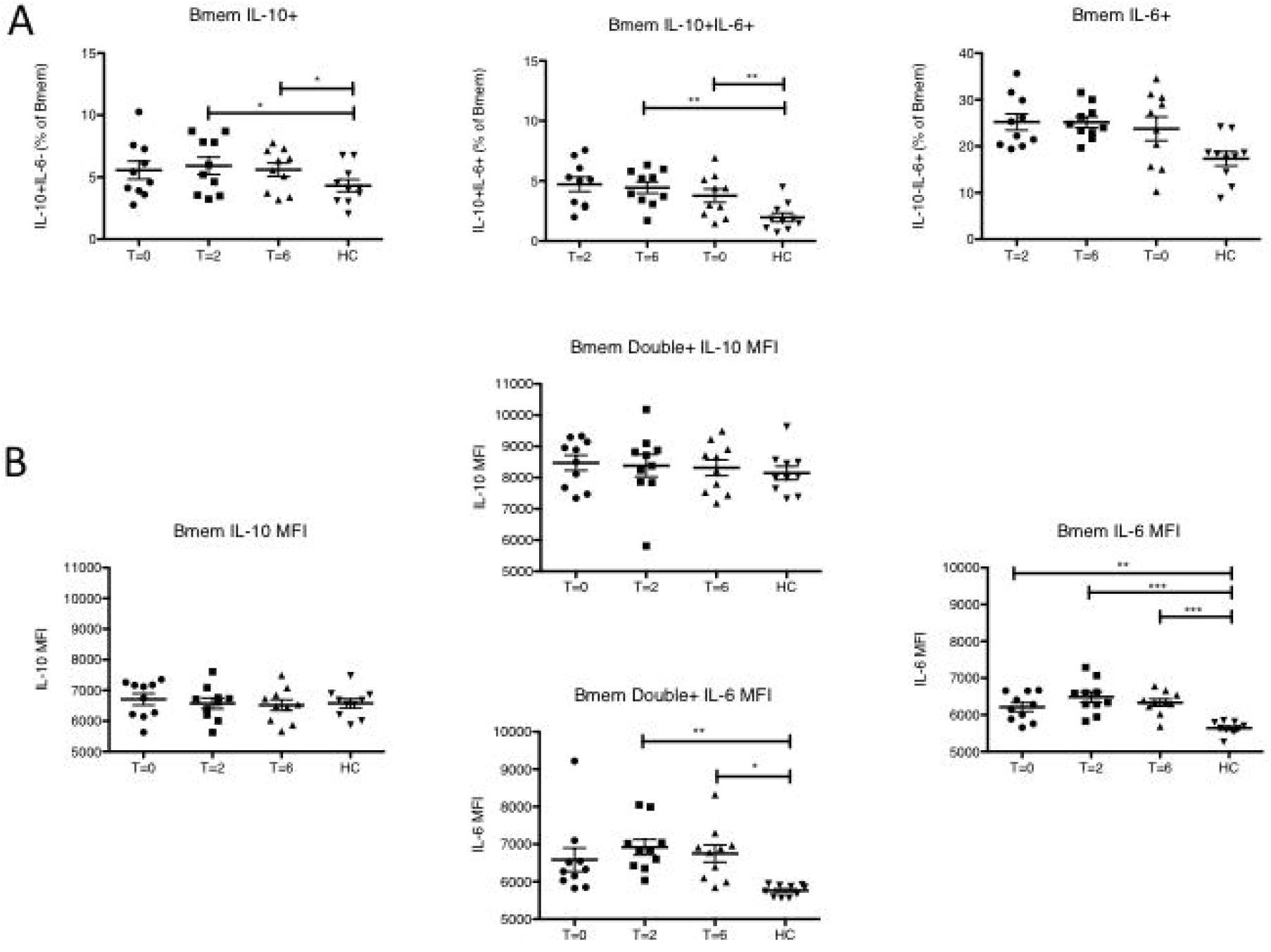
IL-6 and IL-10 expression by memory B cells differs in RRMS patients and healthy controls and is modulated with IFN-β1b treatment. Cells were analyzed via flow cytometry as shown in figure 2. Each data point is representative of the average of two replicates. The horizontal bar indicates the mean frequency for each time point, and error bars represent the standard error of the mean. Data analyzed by one-way ANOVA with Tukey’s multiple comparison post-test. * p<0.05, **p<0.01, ***p<0.001. (A) Frequency of cytokine positive cells memory B cells. (B) Geometric mean fluorescence intensity of cytokine positive cells.

Figure 4B presents the level of expression as measured by geometric mean fluorescence intensity in memory B cells for healthy controls and patients at baseline and two and six months following treatment. In IL-10/IL-6 double positive cells, IL-6 levels are significantly higher at baseline and 2 and 6 months following treatment initiation compared with healthy controls, and IL-10 levels are higher than for single IL-10 positive memory B cells. Expression levels of IL-6 in IL-6 single-positive cells are significantly higher than healthy controls at 2 and 6 months following initiation of IFN-β1b.

### Frequency of IL-10 and IL-6 expression by B1 B cells

The frequency of IL-10 and IL-6 expressing B1 B cells (CD27+CD43+) for all patient time points and healthy controls can be seen in Figure 5A. MS patients treated with IFN-β1b had a significantly higher frequency of IL-10 and IL-6 double producing B cells than healthy controls at both 2 months and 6 months following initiation of treatment (p<0.05). Interestingly, MS patients had a significantly higher level of IL-6 producing B1 B cells at baseline versus healthy controls (p<0.01), and this significant difference continued throughout treatment. Unlike naïve and memory B cells, the frequency of IL-6-secreting B1 cells is reduced by half for both normals and patients.

**Figure 5.**
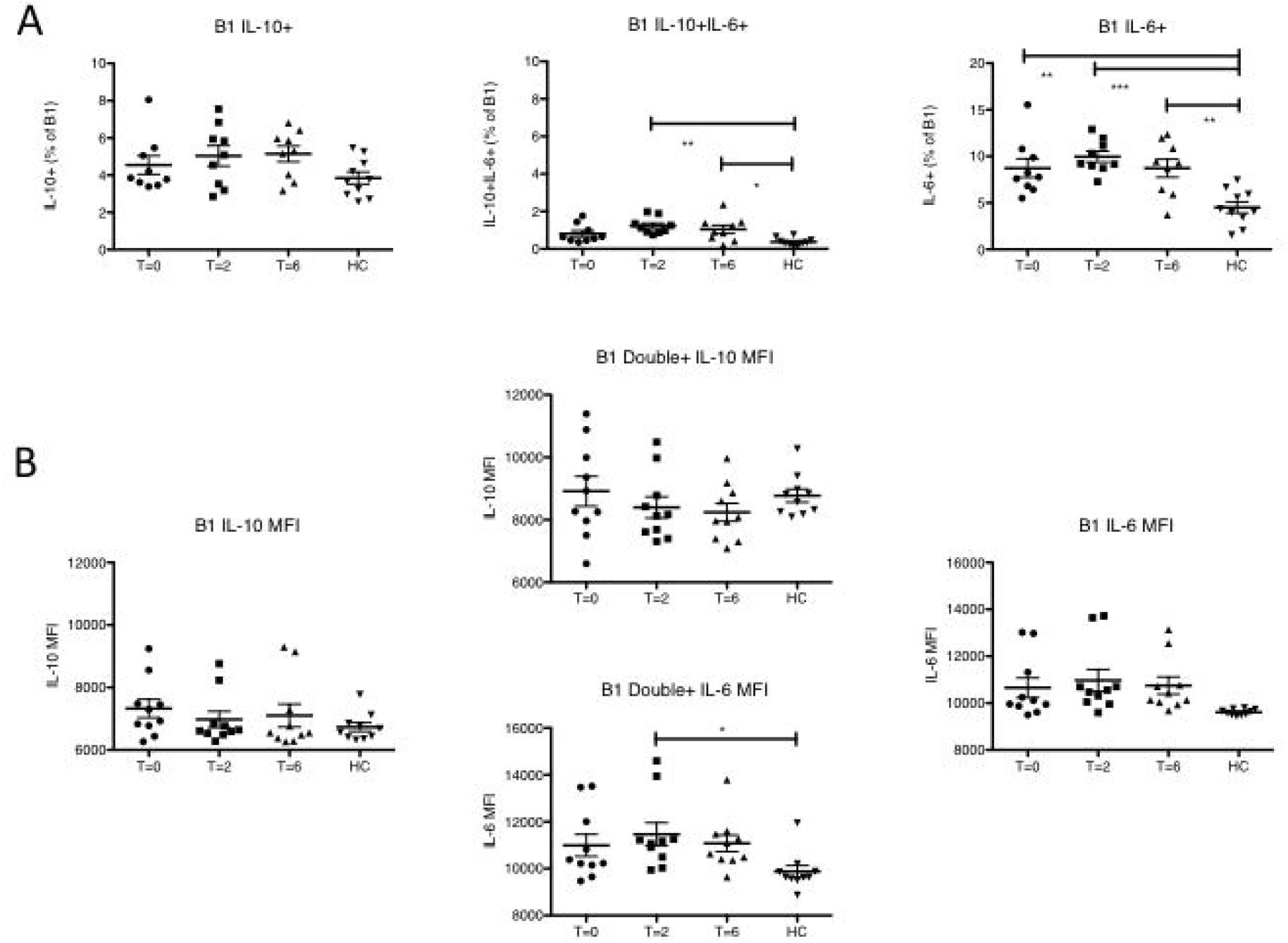
IL-6 and IL-10 expression by B1 B cells differs in RRMS patients and healthy controls and is modulated with IFN-β1b treatment. Cells were analyzed via flow cytometry as shown in figure 2. Each data point is representative of the average of two replicates. The horizontal bar indicates the mean frequency for each time point, and error bars represent the standard error of the mean. Data analyzed by one-way ANOVA with Tukey’s multiple comparison post-test. * p<0.05, **p<0.01, ***p<0.001. (A) Frequency of cytokine positive cells B1 B cells. (B) Geometric mean fluorescence intensity of cytokine positive cells.

Figure 5B presents the level of expression as measured by geometric mean fluorescence intensity in B1 B cells for healthy controls and patients at baseline and two and six months following treatment. In IL-10/IL-6 double positive cells, IL-6 levels are significantly higher at baseline and 2 months following treatment initiation compared with healthy controls. In the single positive populations, no significant differences are seen.

### IL-10 and IL-6 cytokine profile differs in CD5+ and CD5-B1 B cells

B1 B cells in mice are divided into two distinct subsets – CD5+ B1a cells and CD5-B1b cells(24). CD5 expression on B1 cells from MS patients and healthy controls was examined to determine whether there was differential cytokine expression in CD5+ and CD5-cells. As shown in Supplemental Figure 2A, at baseline, as well as 2 and 6 months following initiation of treatment, MS patients showed a significantly higher frequency of CD5 expression, with a mean of approximately 90% while healthy control B1 cells showed a mean frequency of approximately 70%, in line with previously reported figures(10).

IL-10 expression following ex vivo stimulation by CD5+ and CD5-B1 B cells is seen in Supplemental Figure 2B. In the CD5+ population, IL-10 expression is significantly lower in MS patients at baseline compared to healthy controls. Following IFN-β1b treatment, this significant reduction disappears. In the CD5-population, a similar pattern appears, with the frequency of IL-10 expression being lower at baseline and 2 months following initiation of treatment with IFN-β1b, with the significant reduction eliminated at 6 months following treatment. Interestingly, the CD5+ B1 cells have higher overall expression of IL-10 when compared with the CD5-population across all time points and healthy controls. Frequencies of IL-10/IL-6 double positive and IL-6 single positive CD5+ or CD5-B1 cells showed no significant differences between MS patients at any time point versus healthy controls.

### RRMS patients show differing patters of cytokine production among B cell subsets compared with healthy controls that shift with IFN-β1b treatment

To directly compare the contribution of different B cell subsets to the total B cell-derived cytokine, we calculated a cytokine index for each of subsets analyzed. This index is the product of the cytokine positive subset frequency as a percentage of total B cells and the mean fluorescent intensity of the cytokine signal from the cytokine positive cells. As shown in Figure 6A, this index reveals that at baseline, RRMS patients have a similar contribution of IL-10 from each B cell subset, while healthy controls have a much higher contribution from memory B cells. Furthermore, following initiation of IFN-β1b treatment, naïve B cells become the predominant producers of IL-10 among all B cells. IL-6 expression (Figure 6B) shows a different pattern – RRMS patients have expression of IL-6 predominantly by naïve B cells that is unchanged with IFN-β1b treatment, while healthy controls show a reduced level of IL-6 production by naïve B cells that is similar to the amount produced by memory B cells, with little expression of IL-6 by B1 B cells.

**Figure 6.**
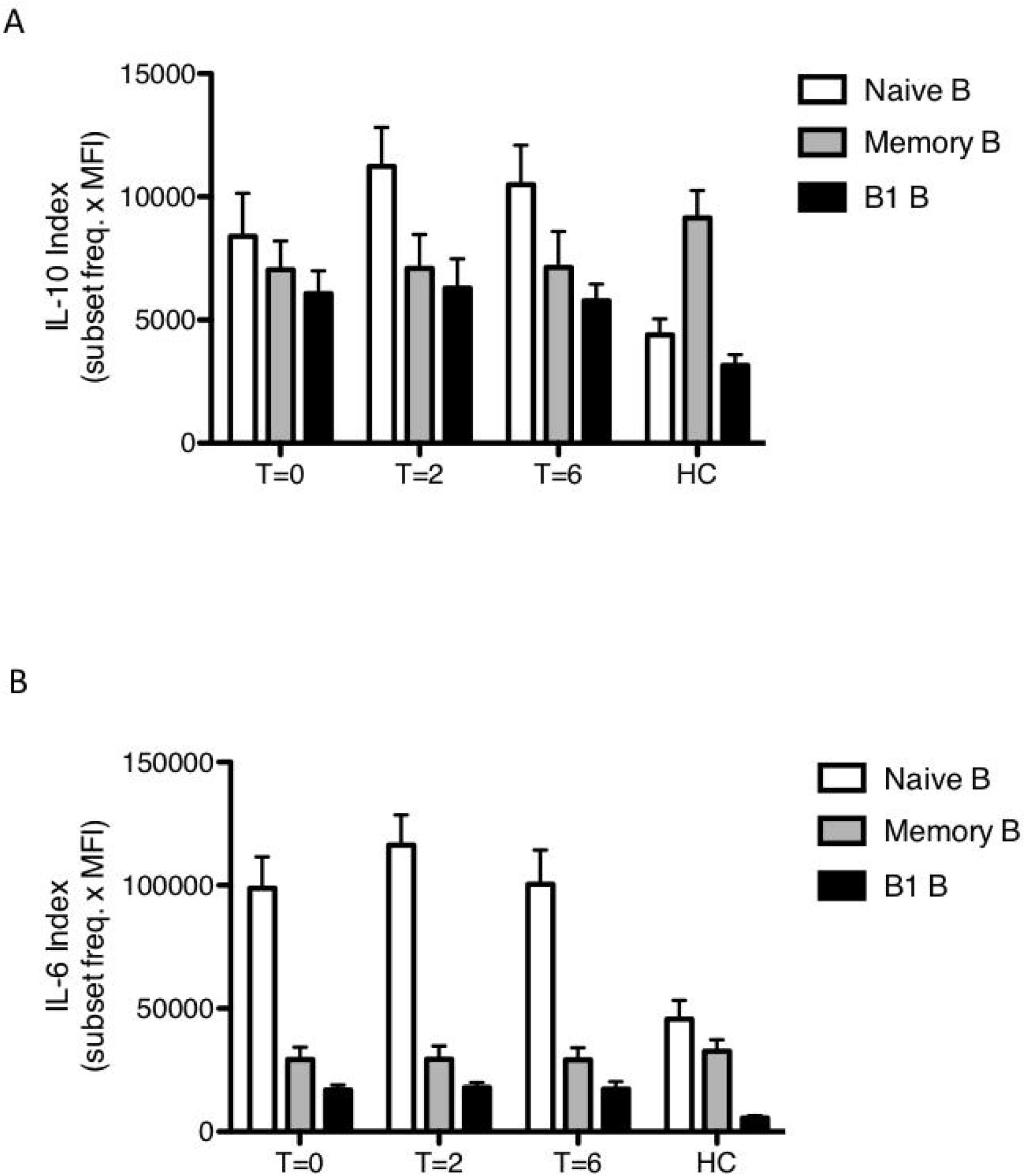
Contribution of IL-10 by B cell subsets is shifted toward naïve B cells in RRMS patients, particularly with IFN-β1b treatment and naïve B cells are the predominant source of B cell-derived IL-6 in RRMS patients. Cells were analyzed via flow cytometry as shown in figure 2. Each bar is representative of the average of 10 patient or healthy control samples. Error bars represent the standard error of the mean. Cytokine index represents the frequency of cytokine positive cells from each B cell subset as a percentage of total B cells multiplied by the mean fluorescence intensity of cytokine positive cells.

### Patients with no or low disease activity following IFN-β1b treatment have significantly different B cell subset frequencies and CD27 expression on B1 cells from patients with moderate or high disease activity

We have demonstrated several differences between RRMS patients and healthy controls, as well as differences following treatment with IFN-β1b in B cell subset composition and cytokine expression. To further determine the effect of IFN-β1b treatment, we divided the RRMS patients into responders to no/low and moderate/high disease activity groups (as defined in the materials and methods) and examined baseline PBMCs for differences. As shown in Figure 7A, patients in the no/low disease activity group have a significantly higher frequency of naïve B cells and a significantly lower frequency of memory B cells than healthy controls following PBMC stimulation. Patients with moderate or high disease activity following treatment had a significantly lower frequency of naïve B cells when compared with the no or low activity group, and a significantly higher frequency of B1 B cells when compared with healthy controls. In addition, as seen in Figure 7B the level of CD27 expression on B1 B cells, but not memory B cells, was significantly higher in the moderate/high group compared with the no/low group. Taken together, these results suggest several differences that could be used as biomarkers to distinguish untreated RRMS patients from healthy controls and, moreover, responders to Betaseron treatment from nonresponders.

**Figure 7.**
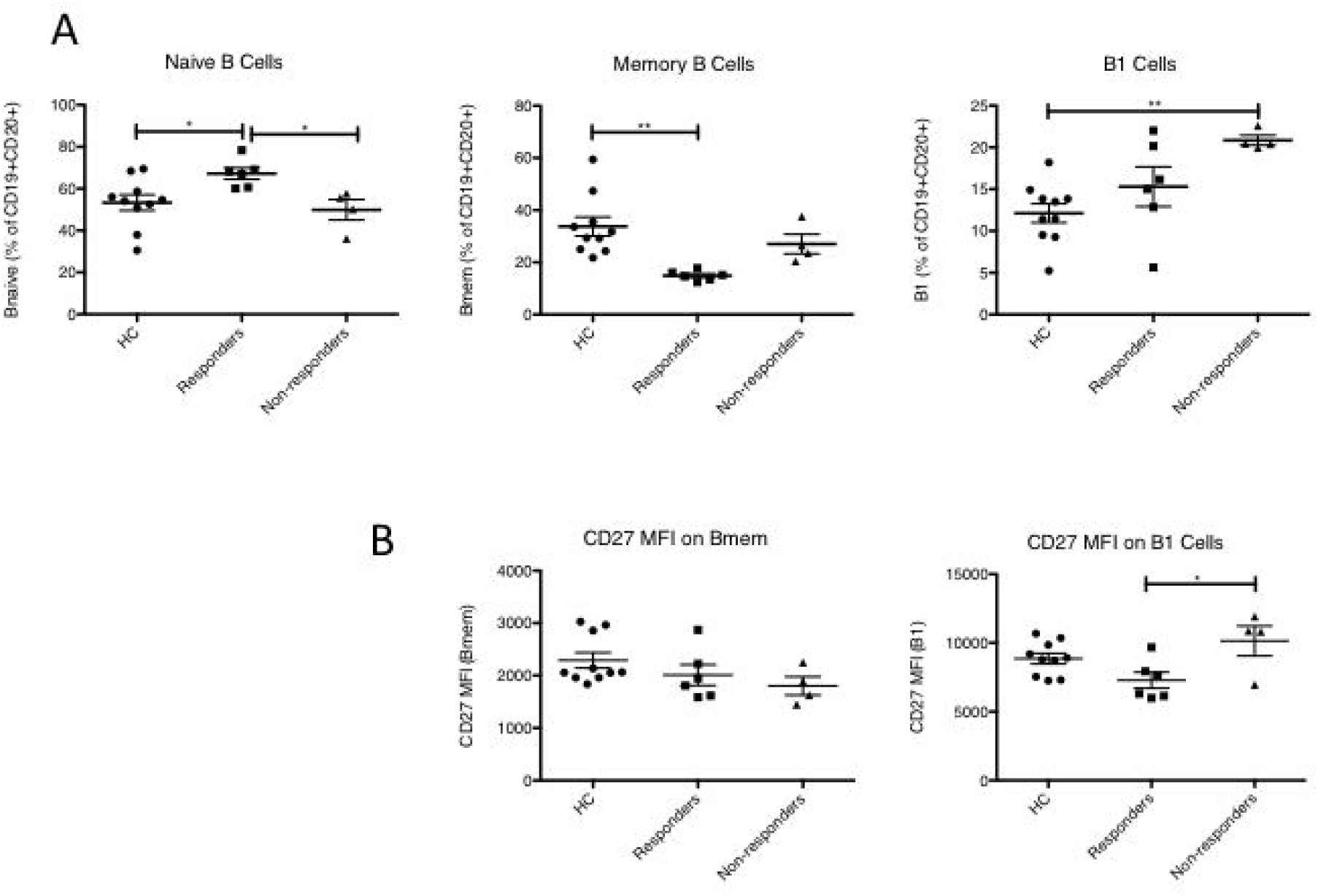
Patients with no/low disease activity and noderate/high disease activity following IFN-β1b treatment have significantly different levels of B cell subsets compared with each other and healthy controls. Cells were analyzed via flow cytometry as shown in figure 2. Each data point is representative of the average of two replicates. The horizontal bar indicates the mean frequency for each time point, and error bars represent the standard error of the mean. Data analyzed by one-way ANOVA with Tukey’s multiple comparison post-test. * p<0.05, **p<0.01. (A) Subset differences between healthy controls (HC), no/low and moderate/high disease activity. (B) Geometric mean fluorescent intensity of CD27 on Memory B Cells and B1 B cells in healthy controls (HC) no/low and moderate/high disease activity.

## Discussion

While multiple sclerosis has been historically thought of as a T cell mediated disorder, B cells are increasingly being appreciated as more central to the pathology of the disease. Oligoclonal antibody bands in the CSF have long been considered a hallmark of the disease, and their source is ectopic germinal centers found in the CNS (25, 26) (27). B cell depleting anti-CD20 therapy has been shown to be effective at treating RRMS, leading to fewer lesions and clinical relapses (4, 5). B cells isolated from RRMS and SPMS patients have been shown to have distinct cytokine secretion profiles compared with those from healthy controls (9). While the role of B cell-depleting mechanisms has been well studied in both human MS and animal models, little is known about the effect of IFN-β1b, a commonly prescribed MS treatment, on the B cell population in MS patients. In this study we demonstrate that RRMS patients compared with healthy controls have notable differences in B cell subsets and cytokine production at baseline as well as showing an effect of IFN-β1b treatment on subset composition and cytokine expression.

Previous studies have looked at the balance between naïve and memory B cells in autoimmune disorders. Until recently, mature circulating B cells have been divided into 2 main subsets: naïve and memory. Naïve B cells have yet to encounter antigen and are defined as CD19+, CD20+ and CD27-. Memory B cells that have seen cognate antigen and represent a pool that respond to secondary infections are defined as CD19+, CD20+ and CD27+. These cell subsets have previously been examined in RRMS patients, with a reduction in memory B cell frequency and an increase in naïve B cells described, with a more profound effect in IFN-β1b treated patients (28). Our data confirms this observation, in that at baseline, RRMS patients had a significantly lower frequency of CD20+CD27+CD43-memory B cells which was further decreased upon initiation of IFN-β1b treatment. The opposite trend was seen with CD20+CD27-naïve B cells: a similar, but slightly higher frequency at baseline that was elevated increased by IFN-β1b treatment.

In the mouse, another subset of mature B cells - B1 B cells - has long been described (29). B1 B cells, which develop in a separate lineage from traditional naïve or memory B cells, produce natural antibody that is cross specific for pathogen and self antigen (30). Natural antibody may be important to modulate the immune response following tissue damage, and MS patients show a differing pattern of expression compared with healthy controls (8). Recently, the human equivalent of the murine B1 cell has been described. These human B1 cells show a similar ability to produce natural antibody, a persistently activated state and a phenotype of CD19+CD20+CD27+CD43+CD70+ (10, 23). We found a significantly increased frequency of these B1 B cells in RRMS patients prior to IFN-β1b treatment, and a significantly decreased frequency with IFN-β1b treatment. Given the elevated level at baseline, and previously reported results showing differential patterns of natural antibody in MS patients, this result suggests that a defect in B1 and/or memory B cell regulation may be involved in the pathology of the disease. A recent study showed a reduction in B1 B cells in newly diagnosed RRMS patients(31). Our study did not focus on recently diagnosed patients – this difference in patient population may account for the differences in subset frequency.

A more recent report describes B cells with the proposed B1 phenotype as pre-plasmablasts, able to differentiate to plasmablasts *in vitro*, that secrete IgG, IgM and IgA and with a gene expression profile between memory B cells and plasmablasts(32). Plasmablasts are effector B cells that have the potential to differentiate into long lived plasma cells, are defined as CD19+CD20-CD27+CD38hi (33). If the cells we have defined as B1 are indeed pre-plasmablasts, the increase seen in this subset in RRMS patients versus healthy controls may be related to the eventual formation of ectopic germinal centers in the CNS and oligoclonal antibody seen in the CSF of RRMS patients. Furthermore, these cells could be precursors to cells that generate anti-self antibody that leads to complement-mediated tissue damage in the CNS. However, the high CD5 expression on the B1 subset that we see - particularly in RRMS patient samples - argues against them being pre-plasmablasts, since plasmablasts do not express CD5.

In addition to being a growth factor for B cells, the cytokine IL-6 is typically a pro-inflammatory cytokine involved in the differentiation of Th17 cells, which are thought to be pathogenic in RRMS. IL-6 produced by B cells has been implicated in the disease process of MS. In the mouse model of MS, experimental autoimmune encephalomyelitis (EAE), B cells from diseased animals produced more IL-6 than healthy controls, and mice with B cells deficient in the ability to make IL-6 showed lower levels of disease (21). In EAE in the marmoset animal model, B cell depletion with anti-CD20 resulted in lower levels of IL-6, as well as other pro-inflammatory cytokines (34). Glatiramer acetate, another approved treatment in MS has also been shown to reduce IL-6 in the mouse model (16). Our results show elevated IL-6 production by naïve and B1 B cells from RRMS patients at baseline compared to healthy controls. Naïve B cells are the predominant B cell subset producing IL-6 in RRMS patients at baseline, and this proportion is unchanged by treatment. This elevated IL-6 expression may be indicative of a dysfunctional B cell population in RRMS patients giving rise to a pro-inflammatory cytokine milieu. Indeed, *in vitro* coculture studies with B and T cells isolated from MS patients show a greater propensity for Th17 skewing compared with cells from healthy controls, providing evidence for this view (35).

The cytokine IL-10 is typically anti-inflammatory, and has been shown to be able to be produced by many different types of leukocytes (13). IL-10 produced by T cells has been shown to be able to suppress Th17 cell proliferation and function (36, 37). IFN-β has been shown to induce T regulatory cells in RRMS patients (38, 39), and these cells can be an important source of IL-10. IL-10 producing B cells, called either B10 or B regulatory cells, have been described in both mice and humans (40, 41). B regulatory cells have been implicated in several EAE studies as being able to modulate initiation or progression of the disease (14, 15, 17). Ex vivo stimulation of B cells from MS patients with CD40 ligand shows a reduced production of IL-10 compared with healthy controls (9). *In vitro* experiments analyzing the effect of IFN-β1b on B cells from both MS patients and healthy controls showed a significant increase in IL-10 production after 24 hours in culture (35). In this study we examined the effect of standard IFN-β1b treatment of RRMS patients on the IL-10 secretion profile of B cells. Overnight *in vitro* stimulation resulted in low frequencies of IL-10 producing B cells in all three subsets of B cells examined, naïve, memory and B1. These cells were present in both untreated RRMS patients and healthy controls, with a trend towards a higher frequency in RRMS patients. The difference seen from the report by Duddy et al (9), - in which MS patients showed a lower amount of IL-10 upon stimulation – may be due to the different choice of cellular stimuli (CD40 ligand previously; CpG in this report) or the fact that the stimulation presented here was on a mixed PBMC population, versus purified B cells. Analyzing the B cells directly via flow cytometry, as in this report, may give a clearer picture of the frequency of these cells to able make IL-10, as a opposed to the indirect method of a supernatant ELISA. All three of the B cell subsets examined are a significant source of the B cell-derived IL-10 at baseline, with naïve B cells becoming a more predominant source with treatment. In contrast, in healthy controls, memory B cells have the highest IL-10 cytokine index.

Consistent with previous descriptions of B regulatory cells, we saw a higher frequency of IL-10 producing B cells in the CD5+ B1 population than in the CD5-population. With IFN-β1b treatment, RRMS patients had significantly more IL-10 production in the memory and naïve B cell subsets than healthy controls, and a trend toward an increase from baseline. This increase in IL-10 from the B cell population may indicate an increase in anti-inflammatory contribution from the B cell population with treatment and may be an important part of the mechanism of IFN-β1b-mediated alleviation of RRMS signs and symptoms.

Several approved first-line treatments for RRMS are available, with similar effects on relapse rate compared with placebo.(42) There are currently no diagnostic tests to assist clinicians in deciding which treatment is advised in a particular patient who is initially presenting with the disease. We have shown significant differences within an RRMS patient population prior to treatment that was correlated with later response to IFN-β1b. Patients with moderate/high disease activity following treatment had significantly higher frequencies of B1 B cells than healthy controls, while patients with no or low disease activity did not, and significantly lower frequencies of naïve B cells. Given that we observed naïve B cells to be the highest producers of IL-6, the fact that patients who showed a better response to treatment had higher levels of these cells may seem counterintuitive. It is possible that the presence of a high level of these IL-6-producing naïve B cells renders the patients more receptive to IFN-β1b treatment, while patients with a lower level of these cells have a different cell type that is driving disease progress. In addition to the differences in cell subset frequency, the level of CD27 on the B1 B cells in the moderate/high disease activity group was significantly higher. CD27 on human B cells has been shown to be a stimulatory receptor leading to an increase in antibody production(43). Thus, the increase in expression in patients less responsive to IFN-β1b may indicate B1 B cells that are more prone to produce auto-antibody. These results may provide a way for clinicians to determine whether IFN-β1b treatment is advised in a particular patient. In order to solidify this finding, further investigations should be performed to determine other markers that may differ in the B cell population between better responders to IFN-β1b and non-responders, and to determine whether serum cytokine level can provide a window into this cellular activity.

The recent evidence arguing for a role of B cells in multiple sclerosis is extensive and compelling. In this study, we demonstrated the effect of a one of the more common treatments for the disease - IFN-β1b - on circulating B cell subsets and the expression of cytokines potentially able to affect the course of MS. RRMS patients had significantly different frequencies of B cell subsets when compared with healthy controls, and IFN-β1b treatment was able to alter the composition of B cell subsets in circulation, decreasing the frequency of memory B cells while increasing the frequency of naïve and B1 B cells. Expression of the pro-inflammatory cytokine IL-6 was higher at baseline in RRMS patients when compared with healthy controls, showing that this population of B cells may be involved in the pathology of the disease. Treatment with IFN-β1b lead to an increase in the anti-inflammatory cytokine IL-10 in RRMS patients compared to healthy controls, indicating that type I interferon may be giving rise to a regulatory B cell population, leading to a reduction in symptoms. Furthermore, when RRMS patients are divided into groups based on disease activity following IFN-β1b treatment, striking differences were seen in B cell subset frequency and CD27 expression. Taken together, this study provides strong evidence that treatment with IFN-β1b can alter the B cell population in RRMS patients, this alteration may be an important part of the mechanism of action of the treatment and treatment efficacy may be able to be predicted by examining the B cell population.

## Data Availability

All data produced in the present study are available upon reasonable request to the authors.

## Acknowledgements

We acknowledge the patients in this study for participating, without whom the work would not have been possible. This work was supported by funding from Bayer Healthcare Pharmeceuticals. Studies were performed in DartLab, the Immunoassay and Flow Cytometry Shared Resource at the Geisel School of Medicine, which receives support from the Norris Cotton Cancer Center and the Dartmouth COBRE Center for Molecular, Cellular and Translational Immunological Research.

## Figure Legends

**Supplemental Figure 1. Frequencies of B cell subsets are stable through a cryopreservation-thaw cycle**. Individual healthy donors were analyzed to assess the stability of B cell subsets on cryopreservation and thawing. Cells were analyzed by flow cytometry (as shown in figure 2) immediately after density gradient separation and following two weeks of cryopreservation. Black bars indicate fresh PBMC; gray bars indicate frozen and thawed PBMC. Error bars represent the range of two technical replicates.

**Supplemental Figure 2. B1 B cells from RRMS patients have higher expression of CD5, and differential expression of IL-10 at baseline**. Cells were analyzed via flow cytometry as shown in figure 2. Each data point is representative of the average of two replicates. The horizontal bar indicates the mean frequency for each time point, and error bars represent the standard error of the mean. Data analyzed by one-way ANOVA with Tukey’s multiple comparison post-test. * p<0.05. (A) CD5 expression on B1 B cells. (B) Frequency of cytokine positive CD5+ and CD5-B1 B cells.

