## Supplementary figures and images for "IFNβ-1b treatment leads to changes in the B cell subset and cytokine secretion profile in patients with relapsing-remitting multiple sclerosis"

### Supplemental Figure 1

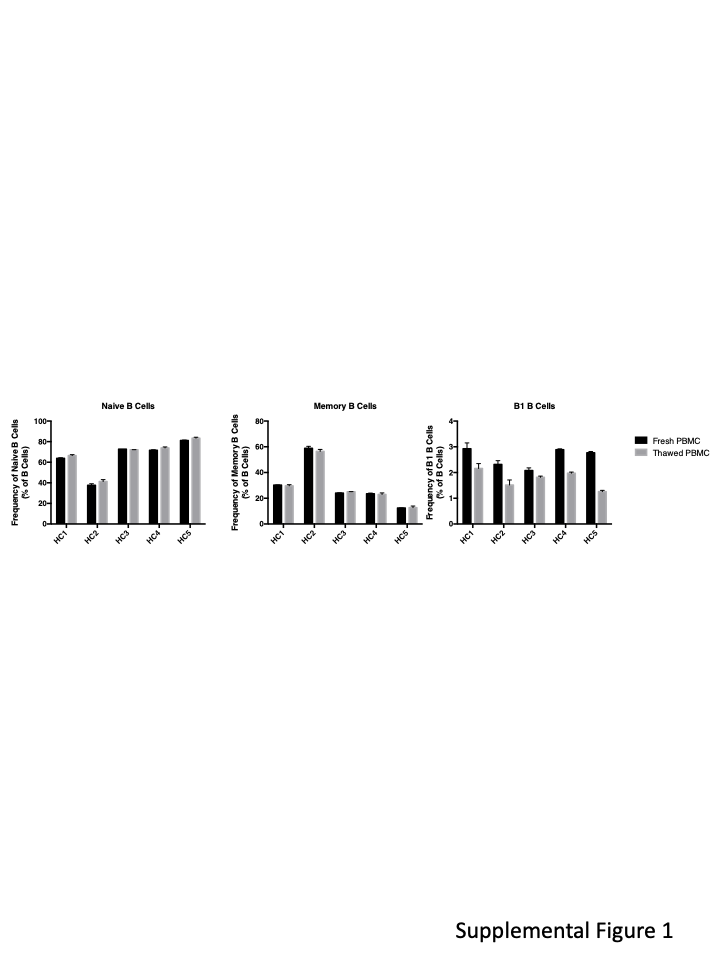

### Supplemental Figure 2

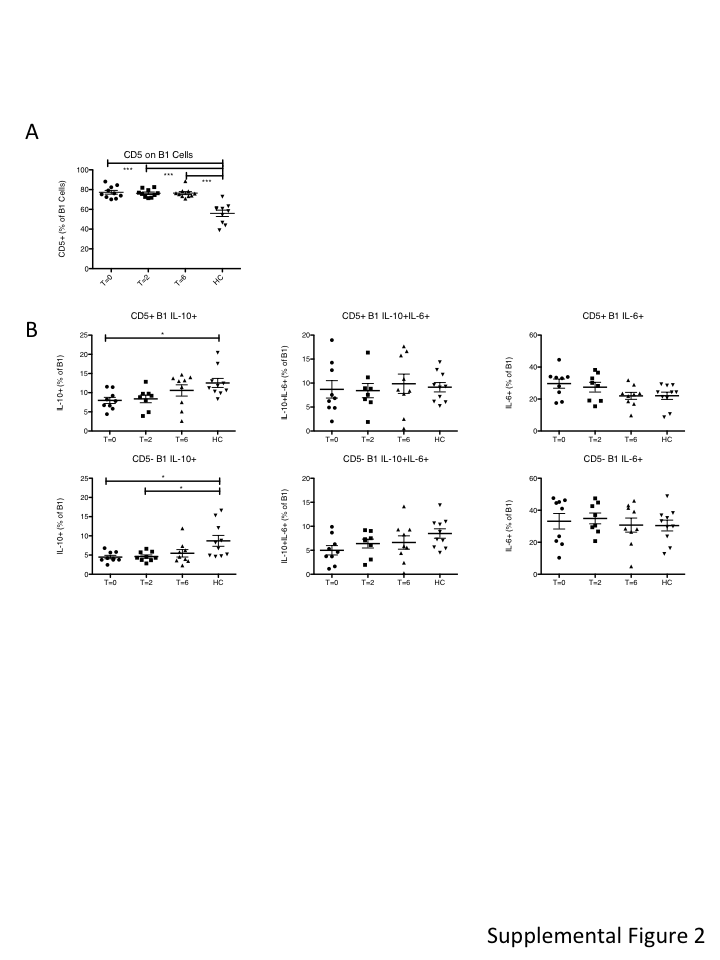
